# Neurochemical and genetic organization of head impact effects on cortical neurophysiology

**DOI:** 10.64898/2026.04.09.26350342

**Authors:** Kevin C. Yu, Laura A. Flashman, Elizabeth M. Davenport, Jillian E. Urban, Srikantan S. Nagarajan, Cormac A. O’Donovan, Kiran K. Solingapuram Sai, Joel D. Stitzel, Joseph A. Maldjian, Alex I. Wiesman, Christopher T. Whitlow

## Abstract

**Purpose:** Previous research has demonstrated effects of head impact exposure on cortical neurophysiology, which may help with understanding variability in clinical sequelae. In separate lines of research, neurochemical and gene transcription markers of vulnerability to traumatic brain injury (TBI) have been established. The purpose of this study was to examine whether these cortical neurochemical and gene transcription gradients are spatially aligned with neurophysiological effects.

**Methods and Materials:** Magnetoencephalography (MEG) data was collected at a total of 278 pre- and post-season timepoints from 91 high school football players across up to four seasons of play. Of the 91 football players, 10 experienced a concussion, and of the remaining 81 non-concussed players, 71 met the criteria for complete imaging and kinematic data, with post-season evaluations less than six weeks after the end of the season. Head impacts were tracked over the course of the season with helmet-mounted sensors. MEG data underwent source-imaging, frequency-transformation, spectral parameterization, and linear modeling to examine the effects of concussive and non-concussive head impact exposure on pre-to-post-season changes in rhythmic and arrhythmic neurophysiological activity. To determine clinical effects, parent reported Post-Concussive Symptom Inventory scores related to cognitive symptoms were correlated with cortical neurophysiological changes. Multi-atlas data of neurochemical system densities from *neuromaps* and gene expression from the *Allen Human Brain Atlas* were examined for alignment with head impact-related alterations in neurophysiology via nonparametric spin-tests with autocorrelation-preserving null models (5,000 Hungarian spins; *p*_FDR_ <.05).

**Results:** Concussion-related reductions in cortical excitability (i.e., aperiodic exponent slowing) were aligned with atlas-based norepinephrine transporter (NET) and alpha-4 beta-2 nicotinic receptor (α4β2) densities, and with apolipoprotein E (APOE) and brain-derived neurotrophic factor (BDNF) expression levels. More severe cognitive symptoms associated with concussion-related slowing of aperiodic neurophysiology were also aligned with atlas-based NET and α4β2 receptor densities. Similar changes in cortical excitability related to non-concussive head impact exposure were colocalized with serotonin receptor (5-HT_1A_) density maps and APOE and BDNF expression. Rhythmic alpha activity was reduced by concussion and colocalized with histamine (H3) and mu-opioid (MOR) receptors, among others, as well as with gene transcription atlases of APOE and C-C chemokine receptor 5 (CCR5).

**Conclusions:** These findings extend our previous work to show that the effects of head impact exposure on neurophysiology are strongest in cortical areas with specific neurochemical and genetic profiles that are known to signal vulnerability to traumatic brain injury, and that these spatial alignments are also associated with self-reported symptom severity.

**Clinical Relevance / Application:** Change in cortical excitability, as measured here by MEG, has potential value as a clinical tool for concussion diagnosis and prognosis. We provide genetic and neurochemical contextualization for these changes that may extend their clinical applications, for example to concussion risk assessment and pharmacotherapies.

## Introduction

Among adolescents in the United States, American tackle football is currently the most popular high school sport and is associated with the greatest number of concussions, or mild traumatic brain injuries (mTBIs).^1–3^ Concussions result in complex pathophysiologic cascades that can produce debilitating clinical burden, involving both microscopic and macroscopic changes in the brain ranging from neurometabolic shifts to ventricular enlargement.^3–7^ Moreover, there is a growing literature suggesting that repetitive non-concussive head impact exposure may also lead to alterations to brain structure and function, disruption of neural circuits, and compromised neurobehavioral functions.^8–10^ Adolescence is a period of particular concern for concussion, as it is characterized by rapid neurodevelopmental change in frontal lobe regions^11^, which are known to be especially susceptible to head trauma.^5,12–19^

Myriad neurochemical systems and genetic factors have been associated with concussion and are continually being studied for diagnostic and therapeutic purposes. Concussion initiates a well described neurometabolic cascade^7^, and a growing body of experimental and translational work also implicates perturbation of neuromodulatory systems after head injury, including dopamine (DA)^20–22^, norepinephrine (NE; i.e., noradrenaline)^23,24^, and acetylcholine (ACh)^25,26^ systems. These same systems undergo developmental remodeling across adolescence, with age- and region-dependent changes in receptor distribution and transporter expression for DA pathways^27^, neuronal nicotinic ACh receptors^28^, and NE transporter^29^.

Catecholamine neurotransmitter systems appear particularly sensitive to head impacts. The axonal projections of the mesocortical, nigrostriatal, and mesolimbic DA pathways are vulnerable to shearing forces caused by impact-related acceleration and deceleration.^20^ Repeated concussions can impair wakefulness and NE signaling by injuring neurons of the locus coeruleus-norepinephrine system.^30^ Cholinergic dysfunction is also induced by mTBI, with several studies demonstrating direct injury to cholinergic projections and massive release of ACh following head trauma, presaging, and potentially contributing to, cognitive symptom severity.^31–33^ Within the cholinergic system, nicotinic acetylcholine receptors (nAChRs) have garnered the most interest. Reductions in hippocampal α7 nAChRs following head trauma are associated with diminished working memory.^26,34^ The α4β2 subtype is among the most abundant nAChRs in the brain,^35^ has considerable interplay with dopaminergic pathways^36^, and has been implicated in various neurological disorders including addiction^36^ and Alzheimer’s disease^37^. However, the α4β2 effects of mTBI on neuronal signaling requires further investigation.

In addition to neurochemical alterations, the expression of several other genes has been implicated in neurotrauma, including brain-derived neurotrophic factor (BDNF), apolipoprotein E (APOE), and C-C chemokine receptor 5 (CCR5). BDNF, the most abundant neurotrophin in the brain, is essential for neuronal differentiation, synaptic plasticity, axonal outgrowth, and neuroprotection.^38^ BDNF is thought to play a complex role influencing injury severity and recovery following mTBI in adults. Studies suggest that the BDNF Val66Met polymorphism may influence post-injury recovery, but findings are mixed.^39–42^ Some human studies report poorer neurocognitive outcomes among Met allele carriers compared to Val/Val homozygotes,^43^ whereas other TBI studies report better executive-function recovery in Met carriers.^40^ CCR5 has also been implicated in brain injury pathology as a regulator of post-injury neuroinflammatory signaling^44,45^, with recent evidence for increased microglial CCR5 expression and CCR5-dependent modulation of oxidative stress responses in animal models of mTBI.^46^ Experimental models further suggest that CCR5 signaling, potentially including neuronal CCR5, may influence recovery and plasticity after injury,^47–49^ although evidence in human mTBI remains limited. Emerging studies have found that APOE expression impairs recovery of the blood brain barrier following traumatic brain injury^50–52^ coinciding with poorer clinical outcomes,^53–55^ and has been associated with increased pathogenic tau accumulation in the context of repetitive mTBIs.^56^

Although these neurochemical and genetic targets continue to be investigated via biochemical assays and immunodetection in animal models, there is a growing demand for neuroimaging modalities that can clinically assess the more subtle, and often dynamic, effects of mTBI on the human brain non-invasively. Magnetoencephalography (MEG) is a non-invasive neuroimaging method that can detect neurophysiological changes with millisecond temporal resolution and may hold clinical value as a tool for mTBI diagnosis and prognosis.^12,57–60^ Previous investigations of football and military related mTBIs have demonstrated alterations in brain signaling in the delta (1 – 4 Hz) and gamma (30 – 80 Hz) frequencies.^12,17,59^ Our recent study indicated that these changes may be indicative of a more parsimonious “slowing” of aperiodic cortical signaling, which also relates to the severity of cognitive symptoms.^61^ Studies of human aperiodic neurophysiology have suggested its utility as a proxy of general cortical excitability^62,63^, aligning our findings with previous *in vitro* and animal research on mTBI^64^. Beyond these aperiodic effects, the residual periodic activity in alpha^61^ and beta^60^ frequency bands appear to also be affected by mTBI.

The aim of the current study was to combine our previously-analyzed MEG dataset^61^ with positron emission tomography (PET) atlases of human cortical neurochemical system densities from neuromaps^65^ and gene transcription topographies from the Allen Human Brain Atlas (AHBA)^66^ to examine potential associations between neurochemical and genetic gradients and head impact-related changes in cortical neurophysiology.

## Materials & Methods

### Participants & Biomechanical Data

The Wake Forest University School of Medicine Institutional Review Board approved and reviewed this study. Assent and written informed consent were obtained from each participant and a parent following detailed description of the study, and all protocols complied with the Declaration of Helsinki. Exclusionary criteria for all participants, based on medical history questionnaire, were comprised of past neurological illness or major psychiatric disease, developmental disorders, medications that alter neurophysiology, concussion occurrence in the preceding year, or any contraindications to MRI or MEG scans (e.g., non-removable ferromagnetic implants).

The imaging Telemetry and Kinematic modeLing (iTAKL) study^12,61,67^ follows high school football players in Winston-Salem, North Carolina to study the effects of head impact exposure on the brain. Biomechanical, neuroimaging (i.e., MEG and MRI), and symptom severity data were collected longitudinally from these players over one or more seasons of play from 2012-2019. Summary demographic data for participants can be found in Table 1. The Head Impact Telemetry System (HITS) was used to collect head impact kinematic data for all football players, which wirelessly transmits accelerometer data to a sideline receiver unit with video recording overlooking the field to verify events. Using this data, the Risk Weighted Exposure (RWE) was calculated, estimating the cumulative risk of concussion over the course of the season based on the quantity and severity of impacts.^68^ The combined probability risk function was used to calculate the combined risk of each event, considering the peak linear acceleration and peak rotational acceleration of each event. The calculated risk was summed to derive the RWE_CP_.^68,69^

Of the football players recruited from the 2012 to 2019 seasons, ninety-one players had complete and useable MEG data at both pre- and post-time points for at least one season (all male; mean age = 16.35), with 57 providing useable data for one season (mean age = 16.43), 22 for two seasons (mean age = 16.33), 10 for three seasons (mean age = 15.88), and two for four seasons (mean age = 16.75). This resulted in 139 complete athlete-seasons of repeated measures data across 91 individuals (Figure 1). Across all seasons, 10 of these players were clinically diagnosed with a concussion during the season of play (mean age = 15.99, mean days between post- and pre-season scans = 143.2, mean days between concussion and post-season scans = 80.6). Concussions were identified by a certified athletic trainer who attended all games and team practices. Players with possible concussions were referred to our collaborating concussion clinic for further evaluation and diagnosis by a Sports Medicine physician. Subsequently, participants underwent neuroimaging and neurobehavioral testing where the parental/caregiver Post Concussion Symptom Inventory (PCSI)^70^ was administered. For concussed participants, timepoints following seasons where a concussion occurred were excluded from our analyses.

**Figure 1.**
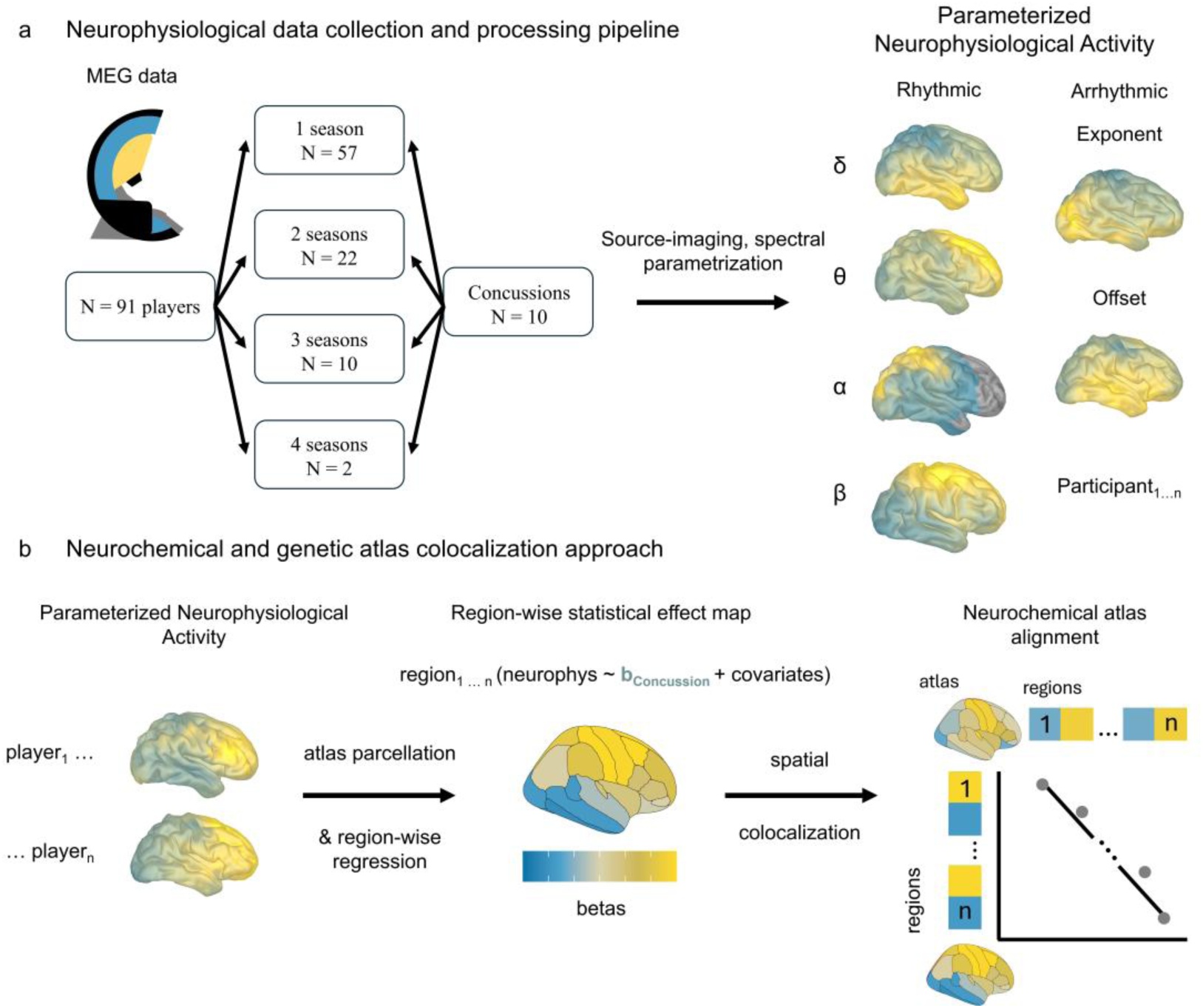
Modeling alignments between neurophysiological alterations, neurochemical systems, and genetic maps in high school football players. (a) Magnetoencephalography data collected from high school football players are preprocessed and source imaged to the cortex. The regional time series are then transformed into the frequency domain and parameterized using *specparam*. (b) The rhythmic and arrhythmic neurophysiological features are averaged over vertices per region of the Desikan-Killiany atlas. The pre-season brain maps are subtracted from the post-season maps to generate maps of neurophysiological change per season and participant. Linear models are fit per region and beta weights extracted to create topographic maps representing the effects of concussion on neurophysiology, which are then tested for alignment with neurochemical and genetic gradients derived from *neuromaps* and the *Allen Human Brain Atlas*, respectively.

Players who did not sustain a concussion were considered for inclusion in the non-concussed analysis using the cumulative impact exposure measure, RWE_CP_. Note that these players were also included as non-concussed participants in the analyses testing effects of diagnosed concussion on neurophysiological activity. Of the 81 football players who did not sustain a concussion during any measured seasons of play, 71 were included who had complete imaging and kinematic data and underwent post-season evaluations less than six weeks after season end (mean age = 16.33, mean days between post- and pre-season scans = 138.62). Football players who underwent post-season evaluations more than six weeks after the season ended were excluded to reduce recovery effects, as well as uncontrolled participation in winter sports or deconditioning exercises.

### MRI Acquisition and Processing

Subjects underwent MRI scanning on a 3T Siemens Skyra MRI scanner with a 32-channel head/neck coil (Siemens Medical, Malvern, PA). T1-weighted anatomical MRI were obtained via a 3D volumetric magnetization prepared rapid gradient echo (MPRAGE) sequence with 0.9 mm isotropic resolution (TR = 1900, TI = 900, TE = 2.93, FA = 9 degrees, 176 slices). Participants had fiducial markers placed on the nasion and bilateral preauricular regions prior to MRI for co-registration with MEG data. Subject anatomical MRIs were segmented and parcellated using *Freesurfer recon-all*^71^ with default settings.

### Magnetoencephalography Data Collection and Analyses

Eight minutes of eyes-open resting-state MEG data^72^ were collected using a 275-channel axial gradiometer CTF Omega 2005 system (VSM MedTech Ltd., Coquitlam, Canada) at a sampling rate of 1200 Hz with participants sitting in an upright position and fixating on a central point. Participants were monitored via real-time audio-video feeds during data acquisition. MEG data was processed using *Brainstorm*.^73^ The MEG data were resampled to 250 Hz, bandpass filtered between 0.5-100 Hz, and notch filtered at 60 Hz (and harmonics). Bad data segments were identified and rejected manually. Independent component analysis (ICA) was used to detect and remove artifacts, including heartbeats, eyeblinks, and muscle artifacts based on their temporal and spatial topographies. The MEG data were coregistered with each subject’s individual structural MRI.^74^ The coregistered data were then source localized using a linearly constrained minimum variance (LCMV) beamformer and an overlapping-spheres head model implemented in *Brainstorm* with default settings, with source orientations unconstrained to the cortical surface at 15,000 locations. Welch’s method (window = 6s; 50% overlap) was used to transform the source-level time series data into the frequency domain. These data were parameterized using *specparam*^75^ (*Brainstorm* MATLAB version; frequency range = 1-40 Hz; Gaussian peak model; peak width limits = 0.5-12 Hz; maximum n peaks = 3; minimum peak height = 3 dB; proximity threshold = 2 standard deviations of the largest peak; fixed aperiodic; no guess weight). To represent the aperiodic components of the neurophysiological power spectrum, the offset and slope of the arrhythmic model fit were extracted per vertex. The aperiodic spectra were then subtracted from the original Welch’s PSD spectra to derive the rhythmic (i.e., aperiodic-corrected) spectra.

The rhythmic and arrhythmic neurophysiological features were subsequently averaged over vertices per each region of the Desikan-Killiany atlas^76^, and the rhythmic spectral data were averaged over canonical frequency bands (delta: 2–4 Hz, theta: 5–7 Hz, alpha: 8–12 Hz, beta: 15–29 Hz). This resulted in six maps of neurophysiological activity for each season and participant: the rhythmic activity in each of the four canonical frequency bands and the arrhythmic activity represented by the aperiodic exponent (i.e., slope) and offset. For each of these features, the pre-season brain maps were subtracted from the post-season maps to generate maps of change in neurophysiological activity per each season and participant. Based on our previous analysis of these data^61^, we focused our hypotheses on aperiodic neurophysiological activity (i.e., the “arrhythmic” slope/exponent) and rhythmic alpha-band (i.e., aperiodic-corrected, 8–12 Hz) signaling.

### Post-Concussive Symptom Inventory

The Post-Concussive Symptom Inventory (PCSI) is a validated 21-item questionnaire that quantifies the severity of a variety of symptoms known to be associated with concussion on a scale of 0 (none) – 6 (worst). Caregiver PCSI data were collected for each participant with diagnosed concussion at time of post-concussion evaluation. Items on the questionnaire are grouped into physical, emotional, and cognitive symptom clusters.^70,77^ As previous developmental literature has demonstrated that concussion sustained at younger ages is associated with worse cognitive function in later adolescence, we focused our hypotheses on the cognitive symptom cluster of the PCSI, which includes: feeling mentally “foggy”, difficulty concentrating, difficulty remembering, getting confused with directions or tasks, answering questions more slowly than usual, and feeling slowed down.^70,77,78^

### Normative Atlases of Neurotransmitter System Density and Genetic Expression

We used the *neuromaps*^65,79^ multi-atlas to represent the spatial distributions of 19 cortical neurotransmitter systems in the healthy human brain: dopamine (D1, D2, DAT), serotonin (5-HT_1A_, 5-HT_1B_, 5-HT_2A_, 5-HT_4_, 5-HT_6_, 5-HTT), acetylcholine (α4β2, M1, VAChT), GABA (GABA_A_), glutamate (NMDA, mGluR5), norepinephrine (NET), histamine (H3), cannabinoids (CB1), and opioids (MOR). For analysis of gene expression levels, we utilized microarray data from the *AHBA*.^66^ Of the available AHBA gene atlases, we investigated the expression of 3 genes that have been previously implicated in the pathogenesis and recovery of head trauma: BDNF^39,40,80^, CCR5^47,48^, and APOE^50–54^.

### Statistics

To represent the region-wise relationship between the occurrence of concussion and pre-to-post season cortical neurophysiological changes, we implemented a linear mixed-effects modeling approach using the *lme4* package^81^ in *R*. A major benefit of this approach is that it allows for the nesting of multiple repeated measures (i.e., seasons) for each participant, while still utilizing all available data. Concussion occurrence was modeled as a categorical variable (i.e., concussion versus no-concussion in a given season), and age at pre-season scan, time between pre-season and post-season scans, and body mass index (BMI) were included as nuisance covariates. The final model took the following form:

### neurophysiological change ∼ concussion + preseason age + time between scans + BMI + (1 | participant)

For each neurophysiological feature of interest, this model was fit per each region of the Desikan-Killiany atlas and the relevant effect statistic (i.e., beta weight) extracted for each region, generating a topographic map of the effects of concussion on pre-to-post-season neurophysiological activity.

We constructed similar region-wise topographic maps of the effects of non-concussive head impact exposure on neurophysiology using linear models. The RWE_CP_ values were log-transformed to account for significant non-normality (Shapiro-Wilk test: W = 0.72, p < 0.001). This linear model incorporating head impact exposure kinematics took the following form:

### neurophysiological change ∼ log(RWE_CP_) + preseason age + time between scans + BMI

Based on our previous findings^61^, we also examined whether changes in aperiodic neurophysiology associated with reported cognitive symptoms of concussion were aligned with cortical neurochemical and gene transcription atlases. For this, we used the following model in the players who experienced a concussion:

### neurophysiological change ∼ PCSI_cognitive_ + preseason age + time between scans + BMI

We tested for spatial alignment between neurochemical/transcription atlases and these effect maps using the *cor.test* function in *R* and non-parametric spin tests with autocorrelation-preserving null models (5,000 Hungarian spins).^82^ Initial models tested for alignments of concussion effects on aperiodic and rhythmic alpha signaling with all neurochemical and gene transcription atlases. Secondary models investigating cognitive symptoms and non-concussive head impact exposures focused on atlases that exhibited significant alignments with concussion effects. Comparisons across all neurochemical and gene transcription alignments per neurophysiological effect topography were corrected using the Benjamini-Hochberg procedure^83^ (*p*_FDR_ < 0.05).

## Results

### Neurochemical and Genetic Influences on Concussion-related Aperiodic Neurophysiological Slowing

We first examined the alignment of concussion-related changes in cortical excitability (i.e., aperiodic slowing) with neurochemical system densities across cortical regions (Desikan-Killiany atlas, n = 68 regions; Figure 2). Concussion-related slowing of aperiodic neurophysiology (i.e., increased exponent) aligned with higher densities of NE transporter (NET; *r* = .43, *p_FDR_* = .001), alpha-4 beta-2 nicotinic receptor (α4β2; *r* = .50, *p_FDR_* = .001), vesicular ACh transporter (VAChT; *r* = .28, *p_FDR_* = .024), 5-hydroxytryptamine receptor 1B (5-HT_1B_; *r* = .30, *p_FDR_* = .024), and histamine H3 receptor (*r* = .28, *p_FDR_* = .028). Concussion-related aperiodic slowing also aligned with lower densities of the dopamine D2 receptor (D2; *r* = -.53, *p_FDR_* = .001), serotonin type 4 receptor (5-HT_4_; *r* = -.44, *p_FDR_* = .004), serotonin 1A receptor (5-HT_1A_; *r* = -.37, *p_FDR_* = .009), serotonin transporter (5-HTT; *r* = -.27, *p_FDR_* = .024), and serotonin 2A receptor (5-HT_2A_; *r* = -.31, *p_FDR_* = .012). Among the considered gene transcription maps, BDNF (*r* = -.47, *p_FDR_* = .001) and APOE (*r* = -.26, *p_FDR_* = .029) expression levels were inversely related to concussion-related aperiodic slowing.

**Figure 2.**
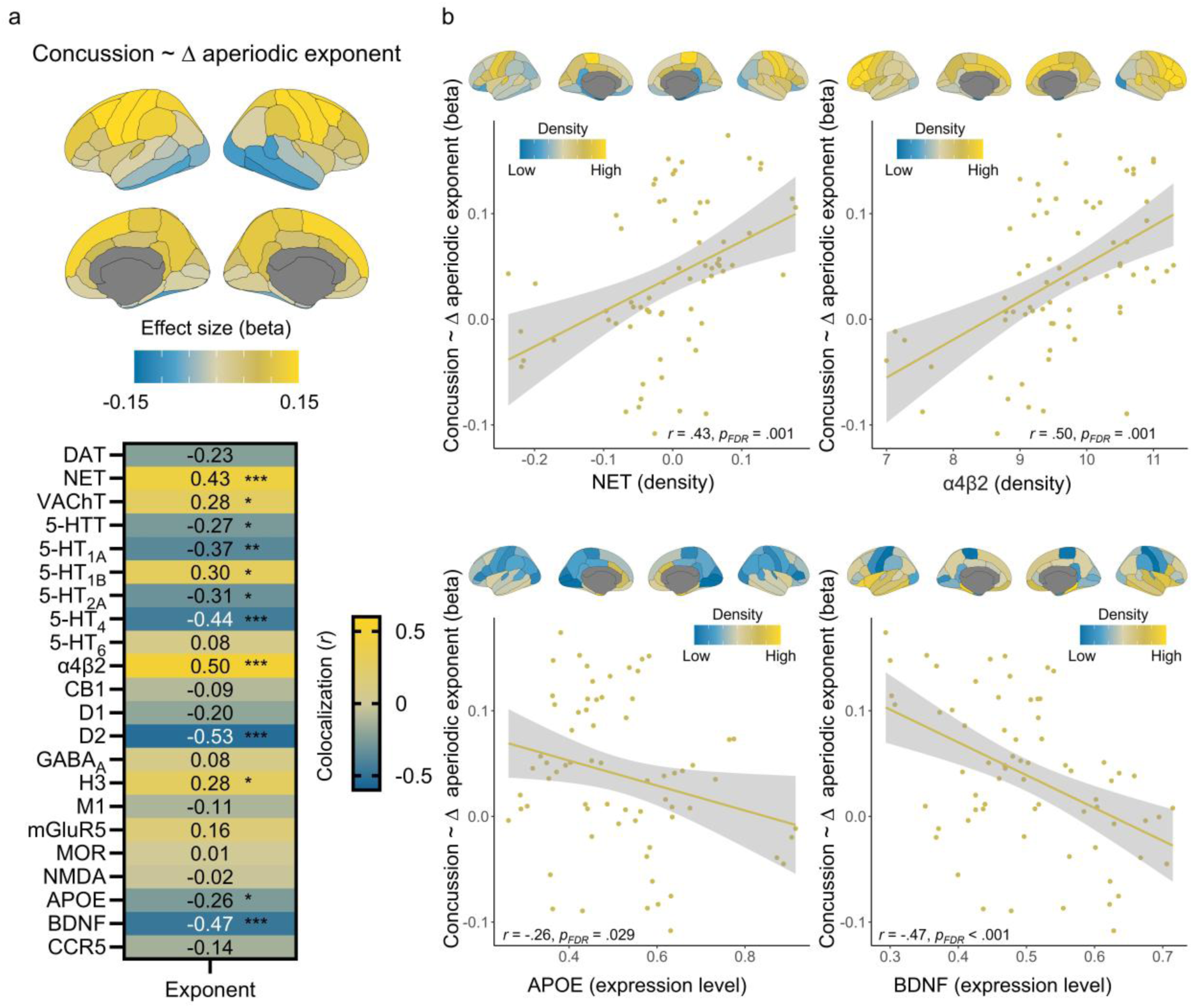
Concussion-related aperiodic neurophysiological slowing is aligned with neurochemical and gene transcription topographies. Concussion effects on the post-minus-pre-season change in aperiodic neurophysiological exponent are displayed in (a) as unthresholded beta weight maps. The heatmap in (a) indicates the strength of the alignment between concussion-related neurophysiological alterations and neurotransmitter and gene transcription topographies, with inferential statistics (Pearson correlation coefficients) overlaid. Scatterplots in (b) show the strongest alignments from (a), with each point representing a Desikan-Killiany cortical region. The cortical maps above each scatterplot portray the topography of each relevant neurochemical and genetic atlas. \*\*\**p*FDR<.005, \*\**p*FDR<.01, \**p*FDR<.05.

### Neurophysiological Changes Associated with Cognitive Symptoms of Concussion Align with Chemoarchitecture

We examined the clinical relevance of the observed concussion-related slowing of aperiodic neurophysiology (Figure 2) within the context of neurochemical and genetic distributions. The association between aperiodic slowing and cognitive symptoms of concussion (i.e., PCSI cognitive scores) in the concussed participants aligned with higher densities of NET (*r* = .56, *p_FDR_* = .001; Figure 3), VAChT (*r* = .64, *p_FDR_* = .001; Figure 3), 5-HT_1B_ (*r* = .35, *p_FDR_* < .001; Figure 3), α4β2 (*r* = .69, *p_FDR_* = .001; Figure 3), and H3 (*r* = .56, *p_FDR_* < .001; Figure 3), as well as reduced densities of 5-HTT (*r* = -.26, *p_FDR_* = .04; Figure 3) and 5-HT_4_ (*r* = -.30, *p_FDR_* = .01; Figure 3).

**Figure 3.**
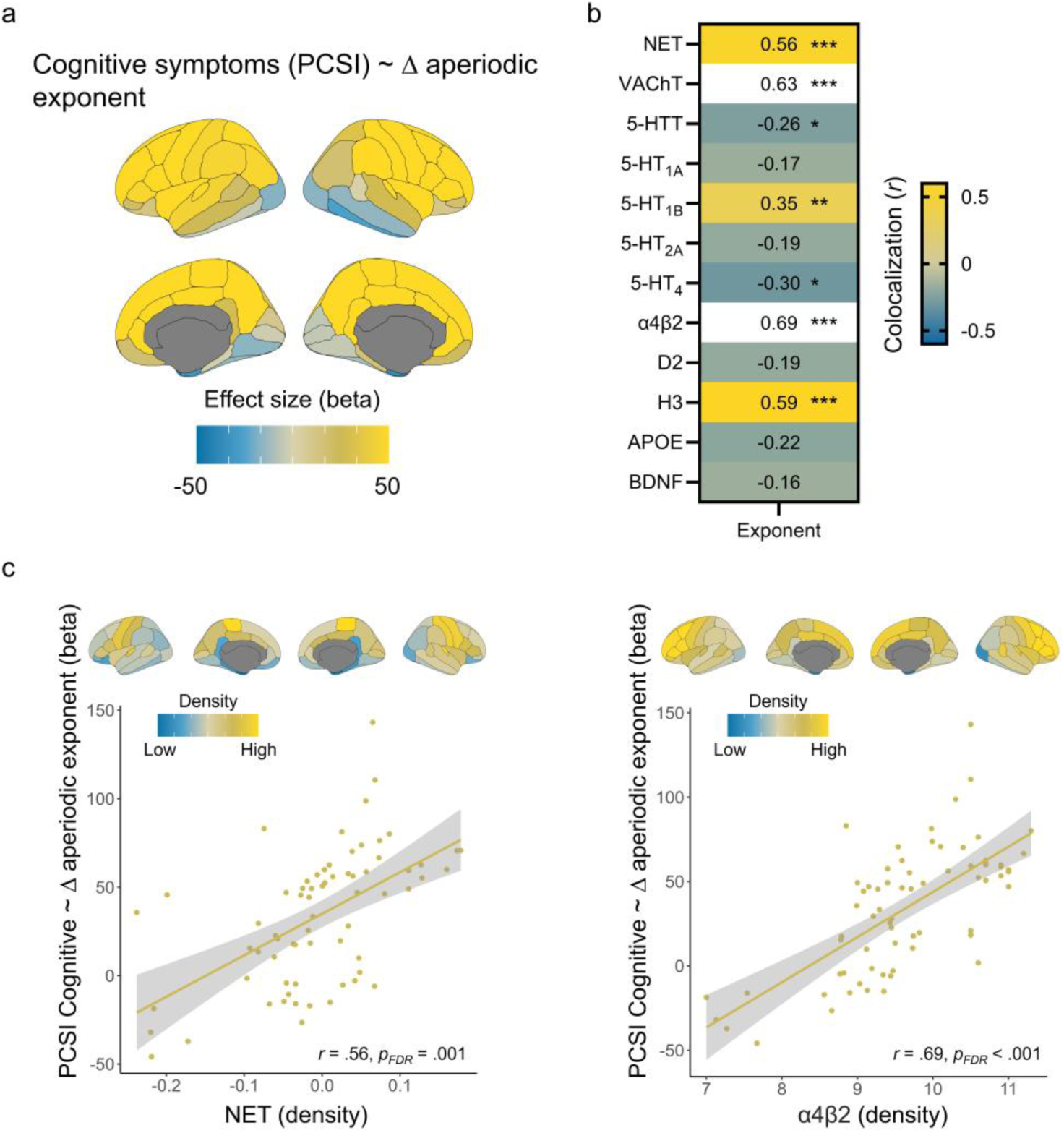
Cognitive symptoms associated with aperiodic slowing align with neurochemical systems. Associations between PCSI cognitive symptom scores and aperiodic slowing in concussion-sensitive regions are displayed in (a) as unthresholded beta weight maps. The heatmap in (b) shows the strength of alignment with neurotransmitter systems and gene transcription. Scatterplots in (c) highlight the significant alignments with NET and α4β2 densities, with each point representing a Desikan-Killiany cortical region. The cortical maps above each scatterplot portray the topography of each relevant neurochemical and genetic atlas. ***pFDR<.005, **pFDR<.01, *pFDR<.05.

### Neurochemical and Genetic Influences on Concussion-Related Changes in Rhythmic Alpha Signaling

We previously observed reductions in rhythmic alpha-band signaling from pre-to-post-season in the concussed group (n = 10)^61^. Akin to our investigations of aperiodic neurophysiology, we sought to explore whether these rhythmic changes aligned with neurotransmitter system densities and genetic transcription (Figure 4). Concussion-related decreases in rhythmic alpha were strongest in regions with high densities of GABA_A_ (*r* = -.47, *p_FDR_* = .003; Figure 4) and low densities of VAChT (*r* = .31, *p_FDR_* = .01; Figure 4), α4β2 (*r* = .34, *p_FDR_* = .02; Figure 4), CB1 (*r* = .40, *p_FDR_* = .008; Figure 4), H3 (*r* = .43, *p_FDR_* = .002; Figure 4), and MOR (*r* = .54, *p_FDR_* = .002; Figure 4). In terms of alignments to genetic expression, concussion-related decreases in alpha rhythms were stronger in regions with lower APOE (*r* = .31, *p_FDR_* = .01; Figure 4) and CCR5 (*r* = .51, *p_FDR_* = .001; Figure 4) expression.

**Figure 4.**
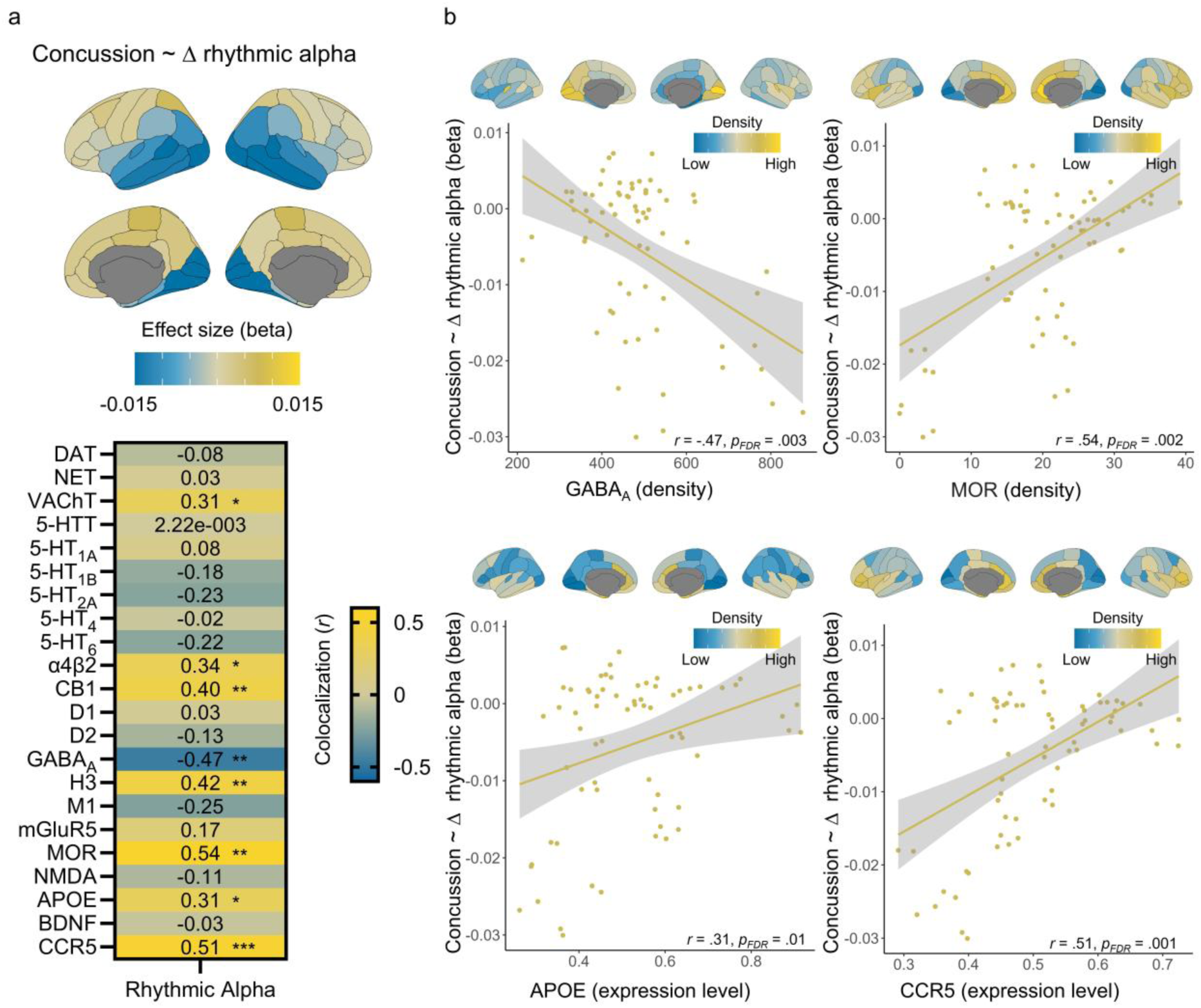
Concussion-related decreases in rhythmic alpha activity are aligned with neurochemical systems and gene transcription. Concussion effects on rhythmic alpha signaling from pre-to-post season are displayed as unthresholded beta weight maps in (a) with a heatmap below indicating significant alignments with neurochemical and gene transcription atlases. The scatterplots to the right (b) highlight significant alignments from (a), with each point representing a Desikan-Killiany cortical region. The cortical maps above each scatterplot portray the topography of each relevant neurochemical and genetic atlas. \*\*\**p*FDR < .005, \*\**p*FDR < .01, \**p*FDR < .05.

### Aperiodic Slowing Associated with Non-concussive Head Impacts is Shaped by Similar Genetic Topographies as in Concussion

In football players who did not receive a diagnosis of concussion but experienced routine head impacts during play (n = 71), we tested colocalizations between aperiodic slowing following cumulative non-concussive head impact exposure (i.e., RWE_CP_) and neurochemical and gene transcription markers. Aperiodic slowing associated with non-concussive head impacts was aligned with 5-HT_1A_ receptor density (*r* = -.36, *p_FDR_* = .034; Figure 5), as well as with APOE (*r* = - .26, *p_FDR_* = .040; Figure 5) and BDNF (*r* = -.24, *p_FDR_* = .040; Figure 5) expression levels. The direction of these effects matched those of diagnosed concussion.

**Figure 5.**
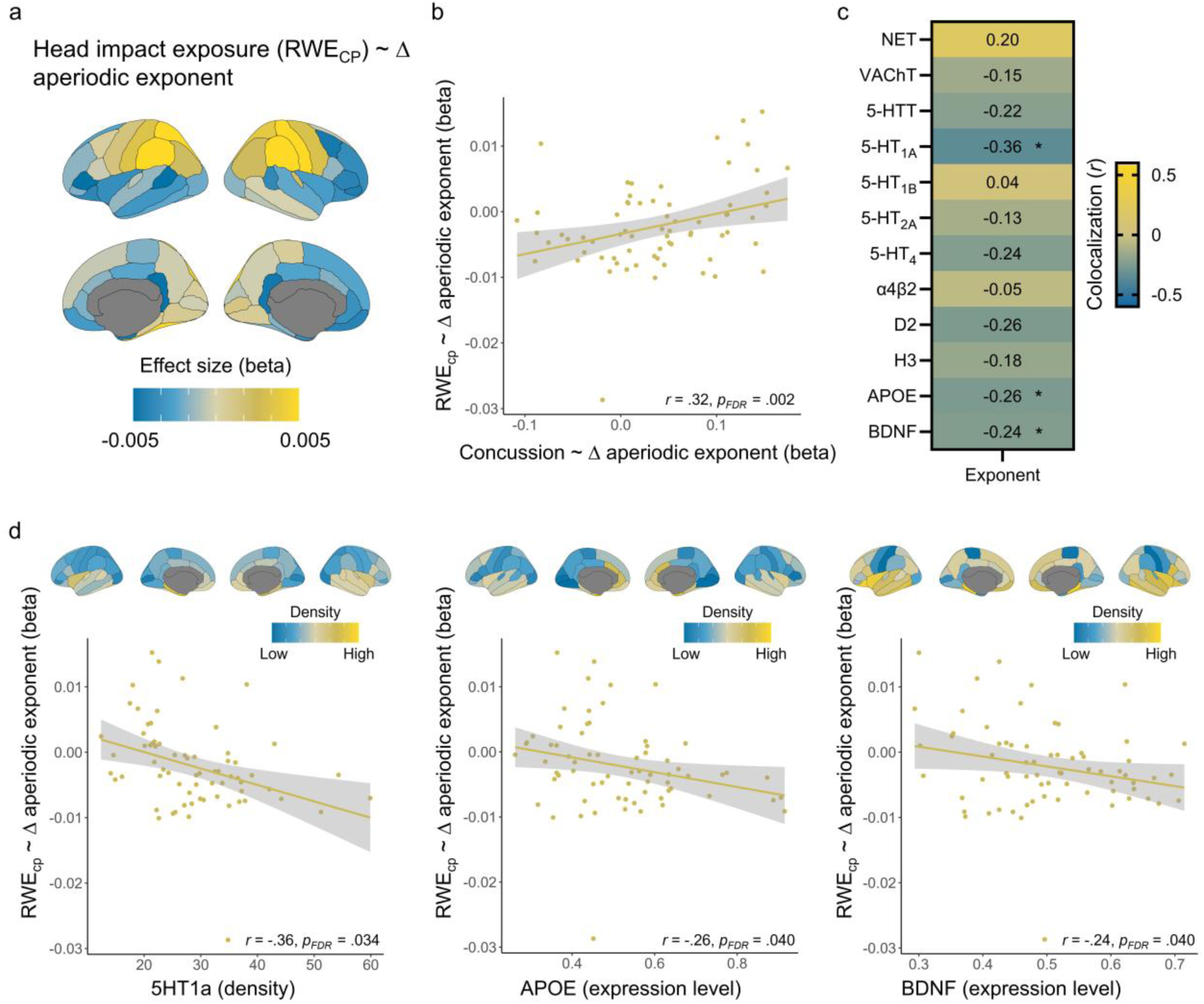
Aperiodic slowing due to non-concussive head impact exposure aligns with serotonergic systems and gene expression. The unthresholded effects of non-concussive head impact exposure on aperiodic neurophysiological activity are shown in (a). The alignment between the concussion effects and non-concussive effects on the aperiodic exponent is displayed in (b). The heatmap in (c) indicates the strength of colocalization between non-concussive neurophysiological alterations and neurochemical and gene transcription atlases. Scatterplots in (d) show the significant alignments from (b), with each point representing a Desikan-Killiany cortical region. The cortical maps above each scatterplot portray the topography of each relevant neurochemical and genetic atlas. \*\*\**p*FDR<.005, \*\**p*FDR<.01, \**p*FDR<.05.

### Changes in Rhythmic Alpha Associated with Non-Concussive Head Impact Exposure Align with Neurochemical and Genetic Gradients

Finally, we wanted to test whether the concussion-related decreases in rhythmic alpha signaling were shaped by similar neurochemical and genetic gradients as those resulting from non-concussive impacts (Figure 6). Of the neurotransmitters and genes that significantly aligned with concussion-related rhythmic alpha changes, we observed that reductions in rhythmic alpha activity related to non-concussive head impact exposure were strongest in regions with low densities/expression of VAChT (*r* = .36, *p_FDR_* = .001; Figure 6), α4β2 (*r* = .35, *p_FDR_* = .005; Figure 6), H3 (*r* = .44, *p_FDR_* = .001; Figure 6), MOR (*r* = .41, *p_FDR_* = .001; Figure 6), APOE (*r* = .24, *p_FDR_* = .029; Figure 6), and CCR5 (*r* = .37, *p_FDR_* = .002; Figure 6).

**Figure 6.**
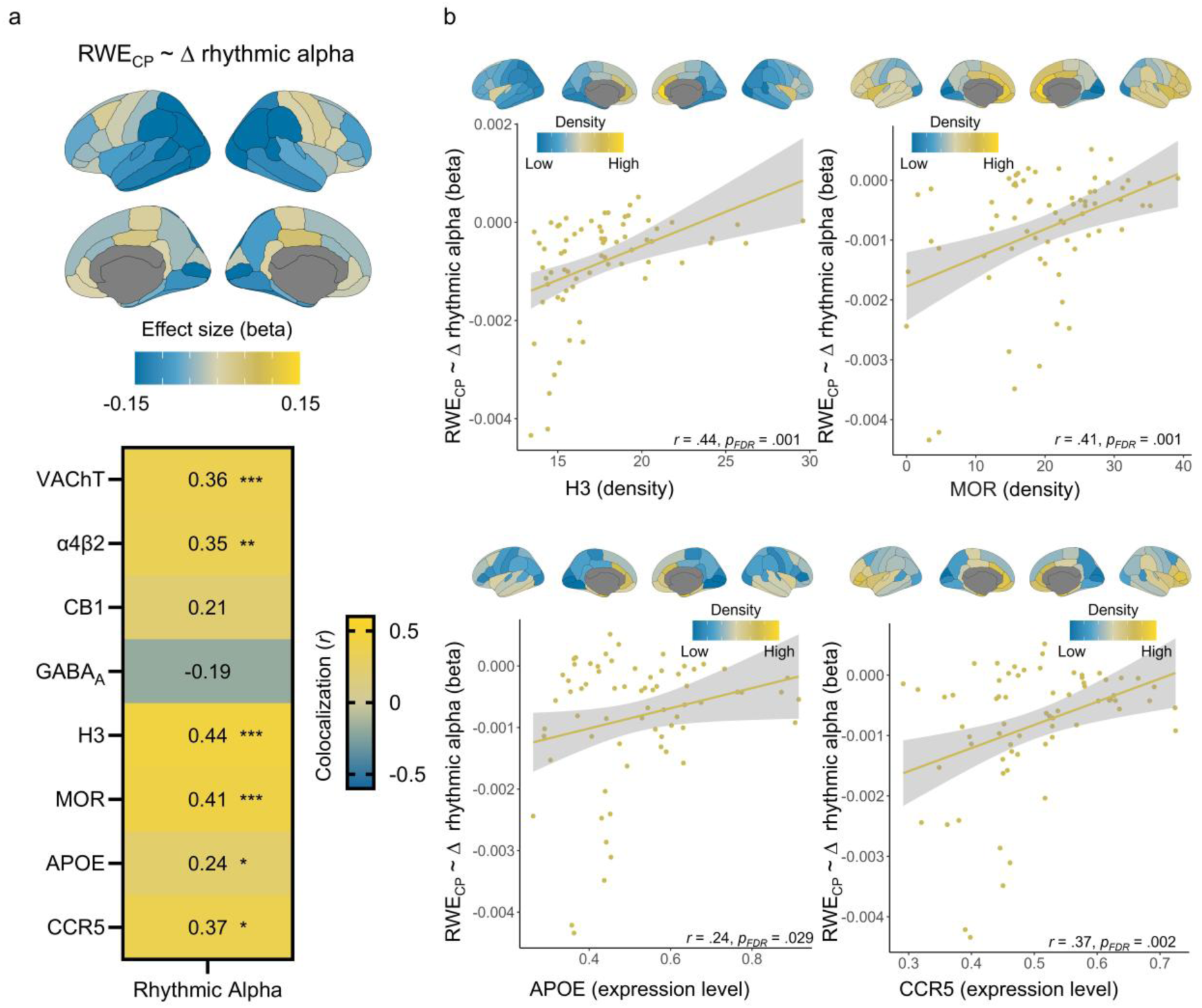
Reduced alpha rhythms due to non-concussive head impact exposure align with neurochemical and gene transcription atlases. The unthresholded effects of head impact exposure on rhythmic alpha activity is shown in (a). Below is a heatmap indicating alignments between these effects and neurochemical and transcription atlases. Scatterplots in (b) highlight significant alignments from (a), with each point representing a Desikan-Killiany cortical region. The cortical maps above each scatterplot portray the topography of each relevant neurochemical and genetic atlas. \*\*\**p*FDR < .005, \*\**p*FDR < .01, \**p*FDR < .05.

## Discussion

These results suggest that both concussive and repetitive non-concussive effects of head impacts on neurophysiology are associated with similar neurochemical and genetic processes, with some notable divergences. We also observe that oscillatory and aperiodic changes in brain signaling induced by concussion align with different neurotransmitter atlases, but consistently with APOE expression, hinting at shared genetic but differential neurochemical bases for effects of concussion on neurophysiology.

We determined that brain regions rich in cholinergic and noradrenergic systems, specifically NET and α4β2, demonstrated the strongest concussion-related slowing of aperiodic brain signaling, indicating reduced cortical excitability. Cholinergic^25,26,31–33^ and noradrenergic^23,24,30^ involvement in the neurochemical cascade of head trauma have been previously studied, though previous investigations have used animal models or been limited to single neurochemical systems in acute phases. In this study, we observed that, relative to other neurochemical systems, atlas-based NET and α4β2 were most strongly related to pre-to-post-season effects of concussion on aperiodic neurophysiology in adolescent football players. Interestingly, the effects of non-concussive impacts did not align as clearly with noradrenergic and cholinergic systems as they did with serotonergic systems. This suggests either that concussion more profoundly impacts brain signaling in these systems, or that greater head impact effects on noradrenergic and cholinergic systems may increase the likelihood of concussion. This latter interpretation is, at least in the case of α4β2, supported by the observation of massive releases of ACh following concussion^31–33^. Moreover, we observed that the aperiodic signaling changes most strongly associated with reported cognitive symptoms occurred in brain regions with high densities of NET and α4β2, indicating further the clinical relevance of aperiodic slowing in cholinergic and noradrenergic systems, and coinciding with previous reports. ^84,85^

The rhythmic alpha signaling decreases following concussion were not aligned with atlas-based NET and α4β2 but rather with GABA_A_ and MOR densities. This is intuitive, as alpha oscillations are thought to be shaped by GABAergic interneurons within thalamo-cortical and cortico-cortical circuits.^86–88^ Additionally, pharmacologic MEG/EEG studies demonstrate that altering GABA_A_ activity with benzodiazepines modulates alpha activity,^89–91^ and PET findings using flumazenil suggest that GABA_A_ receptor availability is altered following TBI.^92^ In contrast, μ-opioid receptors are not typically framed as primary generators of alpha rhythms, but rather as neuromodulatory systems that influence pain, stress, and arousal state, which can secondarily shape rhythmic alpha activity.^93–95^ Together, these results suggest that concussion-related rhythmic alpha changes may be more closely related to inhibitory and state-related neurochemical systems involving GABA_A_ and MOR than to noradrenergic and cholinergic systems.

In addition to neurochemical markers, we also investigated colocalizations with *AHBA* gene transcription atlases.^96^ Based on previous literature, we hypothesized that BDNF would serve a neuroprotective role. In our analyses, we detected an inverse relationship between BDNF expression level and the effects of concussion on changes in the aperiodic exponent, where it appears that brain regions with high BDNF expression were relatively spared from concussive slowing of aperiodic neurophysiology. This pattern was also observed in the non-concussive effects on aperiodic signaling changes, suggesting that BDNF may be protective against both non-concussive and concussive head impacts. These results also appear to indicate that neurochemical systems may play more of a dynamic role in shaping the symptomatic outcomes of head impacts as opposed to the static role of genetic expression patterns.

We did not observe significant alignments between BDNF expression and rhythmic alpha decreases following head impacts. Instead, reductions in rhythmic alpha activity were strongest in cortical regions characterized by lower CCR5 expression. While BDNF has been implicated in neuronal differentiation, synaptic plasticity, and axonal growth^38^, CCR5 has been purported to be a key regulator of neuroinflammation via microglia and neurons^44,45^. Together, these findings suggest a genetic distinction between the effects of concussion on rhythmic versus arrhythmic signalling, whereby rhythmic alpha effects are shaped by the expression of the inflammation-mediating CCR5 while the arrhythmic effects are more impacted by expression of the protective BDNF. In contrast, both rhythmic and arrhythmic neurophysiological effects of head impact exposure were consistently strongest in cortical regions characterized by low normative APOE expression. Although APOE is often discussed in the context of adverse long-term sequelae, especially in relation to the ε4 allele and its importance in Alzheimer’s,^97^ its signaling also exerts well described acute immunomodulatory and neuroprotective effects in experimental models of TBI, including regulation of secondary inflammatory cascades, membrane repair, and blood-brain barrier stabilization.^50–52^ Consistent with this interpretation, multiple preclinical studies have demonstrated that pharmacologic enhancement of apoE-related pathways via mimetic peptides reduces neuroinflammation and improves functional recovery following experimental TBI, including in models of repeated mTBI,^53,98,99^ supporting a protective role for apoE signaling on subacute post-injury timescales.

Our analysis was limited by a relatively small sample of individuals who experienced a concussion during a season of high school football. However, our repeated measures design (i.e., both from pre-to-post season and across seasons within an individual) partially ameliorates this concern by reducing unrelated variance in our signals of interest, as does our secondary analysis demonstrating similar effects of non-concussed head impacts in a much larger sample. The inclusion of only male participants is also a limitation and can be addressed in future studies including female athletes as well as mixed-sex impact sports. Limitations of the neuromaps and AHBA atlases should also be considered, as both resources are derived from adult donor brains, which may not fully capture developmental effects or injury-related plasticity in adolescent populations. However, previous investigations have suggested that the topographies of these systems are relatively stable with age,^100^ and we feel that colocalization of concussion effects with brain areas that are neurochemically-rich in non-concussed individuals still provides useful information, despite limiting interpretations of directionality. Additionally, data collection for this study primarily occurred prior to 2019, when helmet-mounted sensors were used more widely across the field of head acceleration measurement. Helmet-mounted sensor systems have limited coupling compared to newer mouthpiece-based sensor systems but still offer valuable insights into the head impact exposure of athletes. Lastly, this study was limited by the inclusion of only football players from the same league, who likely share similar practice styles and spatial distributions of head impacts. As a result, the observed spatial correspondence between neurophysiological changes and neurochemical or genetic gradients may, in part, reflect underlying biomechanics of head impact exposure rather than neurochemical organization alone. While our findings suggest a relationship between neurophysiological changes and underlying neurochemical and genetic gradients, we cannot fully disentangle these effects from the spatial distribution of biomechanical forces. Future studies incorporating more heterogeneous cohorts and direct measures of regional head impact exposure will be imperative for clarifying the relative contributions of biomechanics and neurobiology, as well as their potential interaction.

Despite these limitations, we have identified genetic and neurochemical systems that differentially shape concussive and non-concussive effects on rhythmic and arrhythmic neurophysiology. We anticipate that these findings will inspire additional targeted studies to not only better understand variability in symptomatic outcomes following head impact exposure, but also the mechanisms of action for the neurochemical markers and gene expressions involved. Identification of how these systems relate to neurophysiological changes in head trauma may inform development of potential pharmacotherapies and treatments, as well as provide guidance for adolescents in contact sports, and illustrate how non-invasive neurophysiology can be used to contextualize injury effects within underlying molecular brain architecture.

## Data Availability

All data produced in the present study are available upon reasonable request to the authors

## Acknowledgments

This work was supported by the National Institutes of Health (NIH) grants R01NS082453 and R01NS091602 to CTW, JDS, and JAM; and the Canada Research Chair (CRC-2023-00300) in Neurophysiology of Aging and Neurodegeneration and a Discovery Grant from the Natural Sciences and Engineering Research Council of Canada (RGPIN-2025-04783) to AIW. Additionally, special thanks to Wake Forest University School of Medicine and the Childress Institute for Pediatric Trauma at Wake Forest Baptist Medical Center for the support provided during the study.

